# Evaluation of tooth sensitivity after scaling and root planing treated with photobiomodulation: a randomized controlled split-mouth double-blind clinical protocol

**DOI:** 10.1101/2023.09.28.23296306

**Authors:** María Victoria García Olazabal, Luis Eduardo Pascuali Moya, Rolf Wilhem Consolandich Cirisola, Ana Paula Taboada Sobral, Laura Hermida Bruno, Federico Todeschini Safi, Priscila Larcher Longo, Maria Cristina Chavantes, Ricardo Scarparo Navarro, Cinthya Cosme Gutierrez Duran, Kristianne Porta Santos Fernandes, Raquel Agnelli Mesquita Ferrari, Sandra Kalil Bussadori, Lara Jansiski Motta, Anna Carolina Ratto Tempestini Horliana

**Author notes:** **Corresponding Author:** (ACRTH). Universidade Nove de Julho (UNINOVE), São Paulo, Brazil (María Victoria García Olazabal, Luis Eduardo Pascuali Moya, Rolf Wilhem Consolandich Cirisola, Laura Hermida Bruno, Federico Todeschini Safi, Priscila Larcher Longo, Maria Cristina Chavantes, Cinthya Cosme Gutierrez Duran, Kristianne Porta Santos Fernandes, Raquel Agnelli Mesquita Ferrari, Sandra Kalil Bussadori, Lara Jansiski Motta, Anna Carolina Ratto Tempestini Horliana). Universidad Católica del Uruguay (UCU), Montevideo, Uruguay. School of Dentistry, Universidade Metropolitana de Santos, Santos, Brazil. Universidade São Judas Tadeu (USJT), São Paulo, Brazil. Universidade Brasil, São Paulo, Brazil (Ricardo Scarparo Navarro). These authors contributed equally to this work (María Victoria García Olazabal, Luis Eduardo Pascuali Moya, Rolf Wilhem Consolandich Cirisola, Laura Hermida Bruno, Federico Todeschini Safi, Priscila Larcher Longo, Ricardo Scarparo Navarro, Anna Carolina Ratto Tempestini Horliana). These authors also contributed equally to this work (Ana Paula Taboada Sobral, Maria Cristina Chavantes, Cinthya Cosme Gutierrez Duran, Kristianne Porta Santos Fernandes, Raquel Agnelli Mesquita Ferrari, Sandra Kalil Bussadori, Lara Jansiski Motta). **Data availability Statement**- all data will be available for the readers. **Funding Statement** - The project received a government grant (National Council for Scientific and Technological Development – CNPq- process: 3146682020-9). In this study the funder only will provide support and resources but will not hold ultimate authority over study design, data collection, analysis, report writing, or publication decisions, ensuring scientific integrity and independence. **Ethics approval Statement** - The project received approval from the Research Ethics Committee of Universidad Católica del Uruguay (UCU), (process: 30052022). **Patient consent Statement** – All the participants included signed an Informed consent. The CI document was in writing. **Clinical trial registration:** NCT05946265 (initial release: July 2023). **Dissemination policy** We will disseminate the trial findings to participants, healthcare professionals, and the general public. Comprehensive data, along with results, will be shared through both complete datasets and results databases while adhering to any publication restrictions. Moreover, we are committed to facilitating potential public access to the full protocol, participant-level dataset, and statistical code. These measures emphasize our unwavering dedication to maintaining transparency. **Author contributions:** Conceptualization; Horliana ACRT, Fernandes KPS, Motta LJ, Bussadori SK; Methodology: Horliana ACRT, Mesquita-Ferrari AR, Sobral APT; Project administration; Bussadori SK; Cirisola RWC; Resources Olazabal MVG, Safi FT; Supervision; Fernandes KPS, Bussadori SK, Mesquita-Ferrari AR; Navarro RS; Validation; Motta LJ, Mesquita-Ferrari AR; Bruno LH, Chavantes MC, Duran CCG; Roles/Writing Moya LEP, Cirisola RWC Olazabal MVG; Longo PL.

## Abstract

It is well known that hypersensitivity affects patients recently treated with scaling and root planing. Some studies have demonstrated that photobiomodulation (PBM) can alleviate dentinal hypersensitivity by modulating pain. However, to date, there is no established protocol for its application after scaling and root planning. To evaluate tooth hypersensibility after photobiomodulation in sensitive scaling and root planning treated teeth. Study design: Randomized, controlled, double-blind split-mouth clinical trial. Methods: Forty-four patients with dentin sensibility after non-surgical scaling and root planning (SRP) will be randomly included in 2 groups: Experimental Group: SRP+ Photobiomodulation (PBM) (660nm, 100W, area 0,5cm^2^, 200w/cm^2^, 30 seconds, 3 J, 6J/cm^2^) and Control Group: Scaling and root planning +FBM simulation. After 7 days of scaling and root planning, all patients will be evaluated for hypersensibility. The cutoff of VAS will be 3. These patients will be included in the study. The primary outcome of the study will be the assessment of dentin hypersensitivity after 7 days of RAR measured with the visual analog scale (VAS). Also, it will be assessed the impact of oral health on the participant’s quality of life, with the OHIP-14 questionnaire. The use of analgesics (paracetamol) will be prescribed as needed and the amount of medication will be calculated. These outcomes will be evaluated after 7 days and 1 month of application. If the data are normal, they will be submitted to the ANOVA test – one way. Data will be presented as means ± SD and the p-value will be set to < 0.05.

## Introduction

Periodontitis is a disease associated with subgingival biofilm adhered to tooth surfaces, which generates a chronic inflammatory response that is unable to interrupt the infectious process and causes progressive destruction of the supporting periodontium (Papapanou et al. 2018).

Periodontal therapy for the management of chronic periodontitis requires non-surgical or surgical intervention to establish healthy periodontal conditions (Alpiste 2006). However, active periodontal therapy causes undesirable sequelae, such as gingival recession or exposure of dentinal tubules, which causes dentin hypersensitivity (Romero 2009). In a previous study (Sobral et al., 1992), of 32 patients with dentin hypersensitivity, 41% had received periodontal treatment and of these, 29% had undergone periodontal surgery. Another study (Hastings 2002) reported that dentin hypersensitivity after periodontal surgery can last for months or indefinitely. Pain arising from the exposed root surface is the most common complaint of patients after undergoing periodontal therapy, however, there is little clinical research dedicated to its treatment (Raut 2018, Corona 2003, Hastings 2002, Sobral 1999, Fischer 1992).

To handle hypersensitivity, treatment with lasers has been proposed (Sgolastra 2013). Laser mechanism action at hypersensitivity is not clearly understood, although several theories have been proposed. High-power lasers (e.g. Er: YAG, Nd: YAG and Er, Cr: YSSG) can reduce or obliterate dentinal tubules by the melting effect on the superficial heat production in cementum (Sgolastra et al., 2011). As for low-intensity lasers, photobiomodulation (FBM) in the dental pulp can cause increased cellular metabolic activity of odontoblasts, intensifying the production of tertiary dentin, causing obliteration of dentinal tubules (Sgolastra 2013, Ladalardo 2004).

The advantage of using low-power lasers is that they are safer, do not emit heat, and are more accessible for clinical practitioners (Ladalardo 2004). For these reasons, the objective of this study will be to evaluate tooth sensitivity after photobiomodulation in sensitive scaling and root-planning-treated teeth.

## Material e métodos

This is a randomized, controlled, double-blind split-mouth clinical trial. It was designed according to the SPIRIT Statement (Figure 1). It was submitted and accepted by the Research Ethics Committee (CEP) of the Catholic University of Uruguay (#30052022). The treatments will be carried out at the Periodontology Clinic II, of the Catholic University of Uruguay in the city of Montevideo, Uruguay, from September 2023 to June 2024. The project has been registered on the ClinicalTrials.gov website (NTC05924204) on 06^th^ June 2023. It is not expected to change the protocol, howev,er any important protocol modifications will be communicated to the Ethical Committee and Clinical Trial Registry. This study does not include minors. All the participants provided informed consent (written and verbal). This document was approved by our Ethics Committee. This study does not report a retrospective study of medical records or archived samples. Any future changes to the study will be promptly reported to the CEP and will be disclosed in subsequent publications. A signed copy of the informed consent form will be provided to each participant.

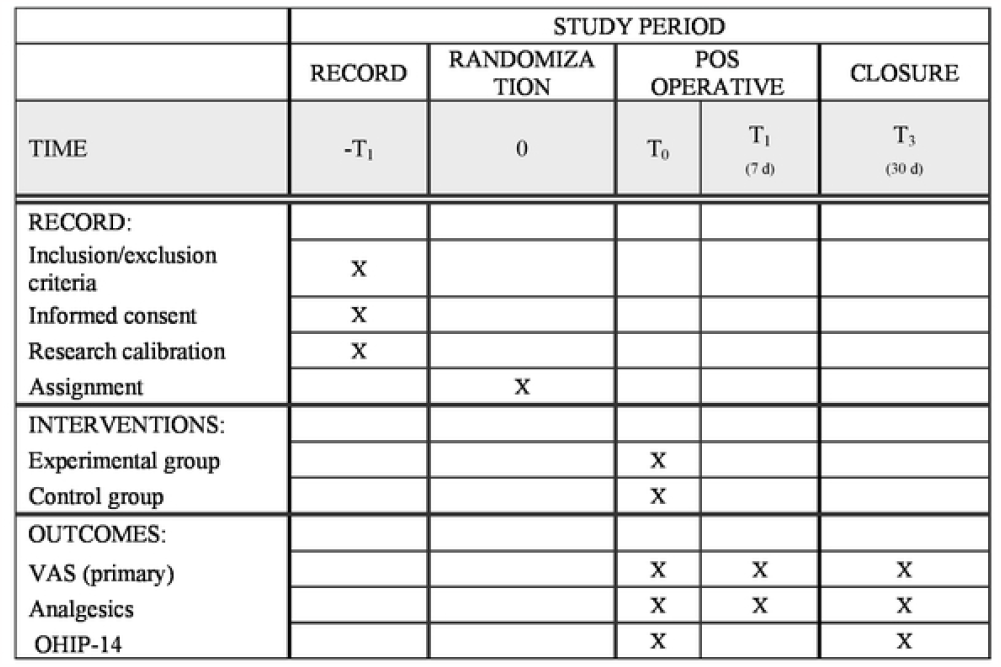

### Sample description

Participants in need of periodontal treatment (scaling and root planning) who seek the service of the Clinic of Periodontics II at the Catholic University of Uruguay with a diagnosis of Periodontitis, stage I, II III, or IV will be recruited by researcher 2. All patients included will receive initial periodontal therapy (educational and preventive interventions and removal of local retentive factors) and the second stage (Scraping and root planning). After a week, they will be probed in the cervical region of all teeth and if there is pain greater than 3 on the VAS, they will be included in the study.

### Inclusion and exclusion criteria

Patients over 18 years of age, both genders, without comorbidities, will be included. Patients who are taking drugs that affect gingival metabolism (cyclosporine, phenytoin, nifedipine), inflammatory medicine (: corticosteroids or non-steroidal anti-inflammatory drugs), or analgesics, pregnant or lactating women, those with photosensitivity history and allergic to paracetamol will be excluded.

### Sample size calculation

The sample size will be 22 patients in each group. This value was calculated to provide a power of 90% (α = 0.05). To determine the necessary n for each group, a sample calculation was performed based on a similar previous study (Ko 2014) which evaluated the outcome pain decrease, measured with the VAS scale. In this study (Ko, 2014) we considered an average of 3 for the experimental group and 5 for the control group. The standard deviation considered for the calculation was ±2, the highest among the groups. A total of 22 patients will be required for each group which totalizes 44 hemiarc (Split mouth).

### Examiner training

The main researcher will be trained to use the visual analogue scale and collect data from the OHIP-14, aiming to maximize the reproducibility of the evaluations.

### Randomization

We will use a random sequence generator website (https://www.sealedenvelope.com/) and select the randomization option of 24 blocks with 2 hemiarcs. The randomization will be implemented on the right side, and the opposite treatment will be applied on the left side. Opaque envelopes will be numbered sequentially and each will contain information about the corresponding group based on the generated order. These envelopes will remain sealed in a secure location until the moment of interventions with FBM or simulation. The randomization process will be managed by an individual who is not directly involved in the study. We have included an additional 6 hemiarches in the sample (44 hemiarches in total) to account for potential losses.

### Blinding

Immediately after periodontal treatment, the researcher responsible for applying the FBM will remove and open one envelope and perform the indicated procedure (FBM or simulation). Only this researcher will know the intervention applied to each patient (Researcher 1). All the other investigators will be blinded to the intervention (researcher 2 - recruitment and researcher 3 - data collection/outcomes) Allocation will not be revealed to participants during the study. I will ensure the confidentiality of personal data for potential and enrolled participants through secure collection, sharing, and maintenance protocols, utilizing a computer with no internet access, password protection, and data encryption stored on a USB drive

### Pre-treatment assessments

Anamnesis, and periodontal examination will be performed. All participants will receive guidance on oral hygiene (HBO), brushing with the Bass technique, specifications regarding the toothbrush (soft bristles and small head), dental floss (traditional Colgate^®^), and scaling and root planning treatment. After one week, periodontal probing will be performed before laser (or simulation application) on treated teeth to assess sensitivity (>3-VAS scale) after root scaling.

All activity performed will be recorded, as well as the amount of medication ingested for pain sensitivity, besides OHIP-14 questionaries.

### Composition of the groups

The 44 hemiarc (22 patients) will be allocated into the groups as follows:

### Experimental Group

RAR + FBM (n=22): The researcher responsible for FBM will remove the randomization envelope and apply Laser Therapy XP (Figure 2) at the points indicated in Figure 3. All dosimetric parameters, details of sessions, and the number of FBM applications are described in Table 1.

**Figure 2.**
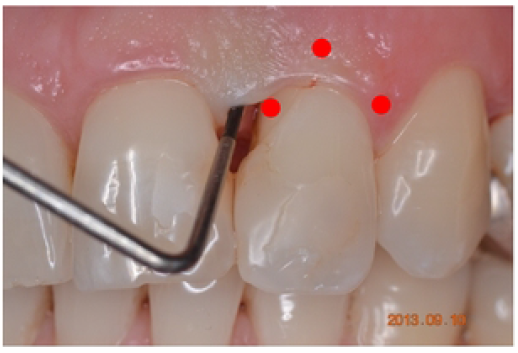
Laser application on the buccal surface (source: adapted from google.com/images)

**Figure 3.**
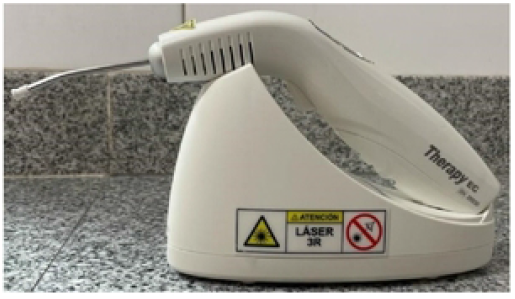
Therapy®EC (DMC) equipment for FBM application.

**Table 1:** Dosimetric Parameters The equipment description, dosimetric parameters, and number of FBM applications are detailed in Table 1.

| Dosimetric parameters | Values/treatment |
| --- | --- |
| Wavelength [nm] | 660 nm |
| Operating mode | Continuous |
| Radiant power [mW] | 100 mW / 0.1 W |
| Irradiance [mW/cm<sup>2</sup>] | 35.385 mW/cm<sup>2</sup> - 35 W/cm<sup>2</sup> |
| Beam area [cm<sup>2</sup>] | 0.002826 cm<sup>2</sup> |
| Exposure time [s] | 30 seconds per point |
| Radiant exposure [J/cm<sup>2</sup>] | 1.061 J/cm<sup>2</sup> |
| Radiant energy [J] | 3 J |
| Number of irradiated points | 6 points per tooth divided into: Vestibular (1 apical, 1 middle, and 1 cervical) and palatal |
| Application technique | In contact, at a 90-degree angle to the surface |
| Number of sessions and frequency | One time only |

### Control Group

RAR + FBM simulation (n=22): Simulation of the use of FBM will be carried out identically to the Experimental group. The person responsible for applying the FBM will simulate irradiation by positioning the devices in the same locations described for the FBM group, however, the laser pointer will be turned off and the sound of the device will be recorded to mimic the use of the equipment, so, participants will not identify to which group they are part of. In this trial, we have no criteria for discontinuing or modifying allocated interventions because no harm is expected with this intervention. The study flowchart presents the details of the project (Figure 4).

**Figure 4.**
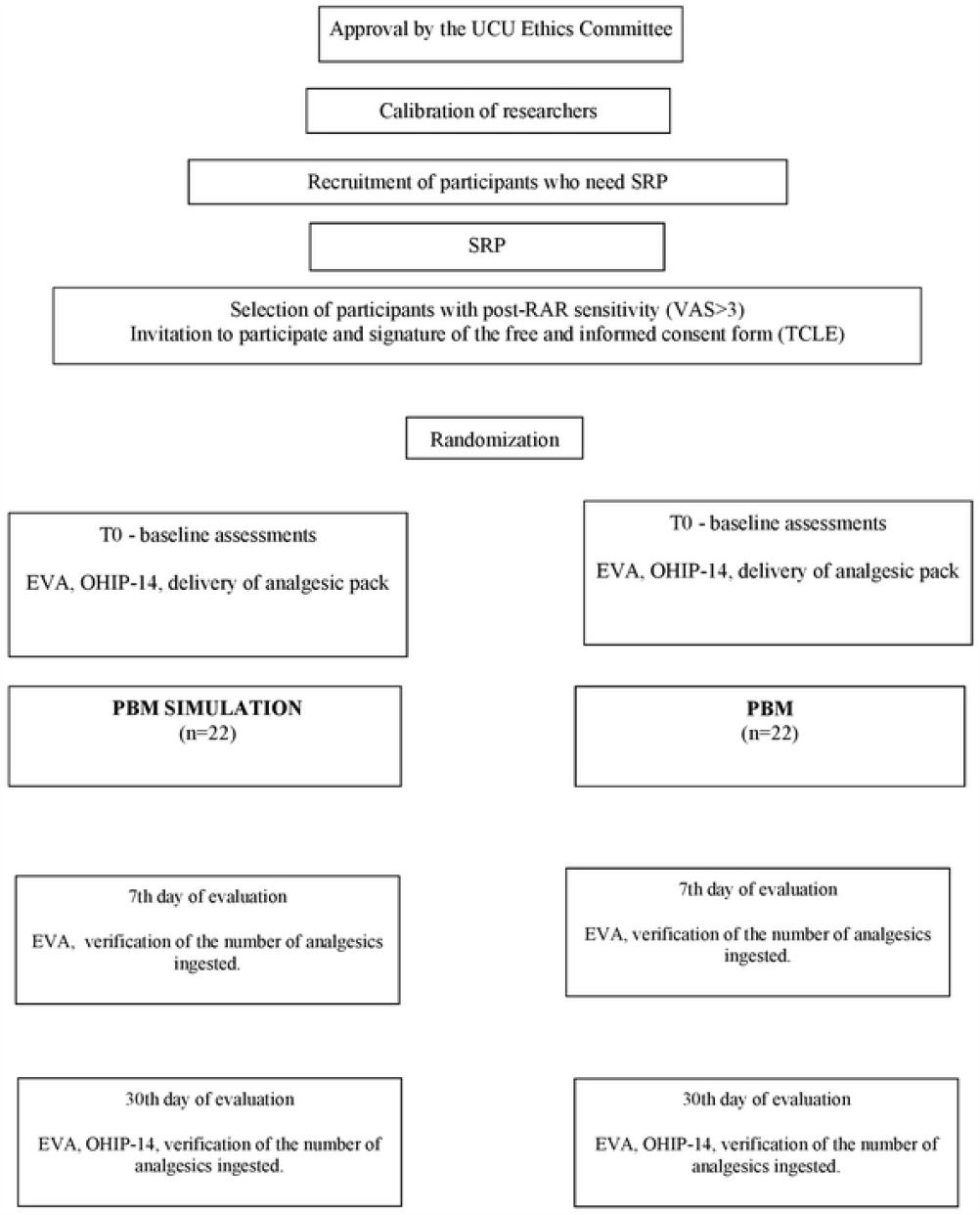
Study flowchart.

### Relevant concomitant care and interventions that are permitted or prohibited during the trial

patients should not take analgesics for dentin hypersensitivity other than paracetamol. Any other mediation (described in the exclusion criteria) will influence the analgesic count in the evaluated period.

### Primary outcome

*Pain -* It will be evaluated through the application of the Visual Analog (VAS) with a 10cm ruler in length of 10 cm, with no mark. One of the extremes is marked “0”, and the other “100” which means respectively “no pain” and “unbearable pain”. This rule will be the same for all participants. Instructions on marking will always be given to the patient by the same operator. Each participant will be instructed to mark with a vertical line the point that best corresponds to the intensity of pain at the time of assessment (Bottega, 2010). Without the presence of the participant, the operator will measure this mark with the same ruler, recording this data on the research clinical record. This analysis will occur on all periods (baseline, 7, and 30 days).

### Secondary outcomes

#### Rescue medication

Another parameter analyzed will be the quantity of analgesics ingested, as proposed by Bauer, 2013. At the beginning of the research, a pack of paracetamol^®^ will be given to each participant (Jóźwiak-Bebenista, 2014) and the use will be prescribed in case of pain. At the end of the experiment, the number of pills will be evaluated as another pain measurement parameter (drug tablet return). To monitor the participant’s adherence they will be asked to take the pack of analgesics to each consultation checking how they will be used. The participant will be oriented to use only paracetamol, not other medicines.

#### Analysis of the impact of oral health on the individual’s quality of life through the application of the OHIP-14 questionnaire (Oral health impact profile questionnaire - Ohip-14)

The Ohip-14 will be used to assess the impact of oral health on the participant’s quality of life (Slade, 1997). To quantify the Likert scale, we will use the additive method – Points will be summed (0-56), with 56 having the greatest impact on quality of life. This is validated and translated into a Portuguese questionnaire (Riva, 2022).

### Any changes in the protocol will be communicated to the Ethics Committee

#### Analysis of results

Initial descriptive analyses will consider all the variables measured in the study, both quantitative (mean and standard deviation) and qualitative (frequencies and percentages). Subsequently, normality analyses will be performed to determine the appropriate statistical tests for each data set and the appropriate statistical tests will be applied for each specific analysis. In all tests, the significance level of 5% of probability or the corresponding p-value will be adopted. All analyses will be performed using the statistical program SPSS for Windows, version 9.1. The data of all patients randomized for the research will be included in the statistical analysis, described, and discussed, as well as the possible adverse effects. Adverse events will be questioned for all participants during the experiment. However interim analyses will not plan because adverse events are not expected in this trial.

## Data Availability

Deidentified research data will be made publicly available when the study is Completed and published.

